# The distress of Iranian adults during the Covid-19 pandemic – More distressed than the Chinese and with different predictors

**DOI:** 10.1101/2020.04.03.20052571

**Authors:** Asghar Afshar Jahanshahi, Maryam Mokhtari Dinani, Abbas Nazarian Madavani, Jizhen Li, Stephen X. Zhang

## Abstract

Early papers on the mental health of the public during the Covid-19 pandemic surveyed participants from China. Outside of China, Iran has emerged as one of the most affected countries with a high death count and rate. The paper presents the first empirical evidence from Iranian adults during the Covid-19 pandemic on their level of distress and its predictors. On March 25–28, 2020, a dire time for Covid-19 in Iran, we surveyed 1058 adults from all 30 provinces in Iran using the Covid-19 Peritraumatic Distress Index (CPDI). The distress level of Iranian adults (mean: 34.54; s.d.: 14.92) was significantly higher (mean difference: 10.9; *t*=22.7; *p*<0.0001; 95% *CI*: 10.0 to 11.8) than that of Chinese adults (mean: 23.65; s.d.: 5.45) as reported in a prior study with the same measure of Covid-19 Peritraumatic Distress Index (CPDI). We also found the predictors of distress in Iran vary from those in China. Our findings that the predictors of distress in Iran vary from those in China suggest the need to study the predictors of mental health in individual countries during the Covid-19 pandemic to effectively identify and screen for those more susceptible to mental health issues.

**Funding:** None

## Introduction

Covid-19 disrupts lives and work and causes psychological distress to the general public^1–4^. The Covid-19 outbreak first triggered public panic and mental health distress in China^2^. Early research has found working adults in more affected areas in China had worse health conditions, more distress and lower life satisfactions^3^. Researchers at Shanghai Mental Health Center developed a Covid-19 Peritraumatic Distress Index (CPDI) to assess the distress level specific to Covid-19^5^. Such research on mental health during the Covid-19 pandemic is critical to identify and screen people psychiatrically based on their distress levels to prioritize mental health services online during the ongoing crisis^2,6^. The identification of those who are more likely to suffer mentally enables more targeted assistance from caregivers and policymakers, especially given the limited resources relative to the scale of the pandemic^1,3^.

One of the countries most affected by Covid-19 is Iran^7^. When we started our survey on March 28, 2020, Iran had one of the highest national counts of Covid-19 confirmed cases (35,408) and deaths (2717), and a mortality rate of 7.6%, as reported by the Iranian government. The figures reported by *BBC Persian* for Iran, however, were much higher. The Covid-19 outbreak in Iran has been compounded by the ongoing decade-long US-led sanctions on Iran. A *Lancet* correspondence noted that “all aspects of prevention, diagnosis, and treatment are directly and indirectly hampered, and the country (Iran) is falling short in combating the crisis. Lack of medical, pharmaceutical, and laboratory equipment such as protective gowns and necessary medication has been scaling up the burden of the epidemic and the number of casualties”^7^. We provide the first empirical evidence on Iranian adults’ level of distress and identify several predictors of their distress under the Covid-19 pandemic to enable early screening for targeting mental health services online.

## Methods

We surveyed Iranian adults aged 18 and above March 25–28, 2020. On those four days, there were 27,017, 29,406, 32,332 and 35,408 total confirmed cases and 2077, 2234, 2378 and 2517 deaths respectively in Iran. The situation was dire. For instance, on March 27, Iranian media reported that about 300 people died due to methanol poisoning in a desperate hope that drinking alcohol kills the virus, even though the Islamic state of Iran forbids the sale of alcohol for drinking. On March 28, prisoners in several prisons were distressed enough that they clashed with guards, set prisons on fire, and somehow escaped.

Given the dire situation in Iran and the lockdown to contain the virus, we delivered the survey online across all 30 provinces in Iran. The survey got the ethics approval (#20200304) at Tsinghua University. We promised the respondents confidentiality and anonymity of their responses.

Participants answered the survey on their gender, age bracket, education level, number of children, whether they had Covid-19, exercise hours per day in the past week, working situation (work from home; work in office, etc.), employment status (employed, unemployed, studying, and retired) and the Covid-19 Peritraumatic Distress Index (CPDI)^5^. CPDI is an index of 24 questions designed to capture the frequency of specific phobias and stress disorders relevant to Covid-19, including anxiety, depression, specific phobias, cognitive change, avoidance and compulsive behavior, physical symptoms and loss of social functioning^5^. CPDI ranges from 0 to 100. CPDI was originally developed in Chinese, and we had the index translated from its English version to Persian. The English and Persian versions of CPDI, as well as its scoring key can be found in the appendix.

## Results

Our online survey received 1058 responses from all 30 provinces in Iran. Table 1 contains the descriptive characteristics of the participants. Of the participants, 53.8% were female, 869 (82.1%) reported being free from the disease, 7 (0.07%) reported having Covid-19, and 182 (17.2%) were unsure of their disease status. In the past week, 30.0%, 4.3% and 1.1% of the participants exercised on average 1, 2 and 3 hours per day respectively, 62.9% of them did not exercise in the past week, and the remaining 1.3% exercised 4 hours or more per day. By employment status, 71.5% were employed, 13.2% were unemployed, 10.5% were studying, and the rest 4.7% were retired. By working situations, 38.6% were working/studying from home, 26.6% were not working/studying due to the outbreak, 18.0% were not working/studying before and during the outbreak, and 16.9% still worked in an office.

**Table 1.** Descriptions of the participants and OLS regression results on CPDI (Covid-19 Peritraumatic Distress Index) (*n*=1058)

|  | Description | Regression parameters |  |  |  |
| --- | --- | --- | --- | --- | --- |
| Variable | Count (%) | Coef. | P | Lower 95% C.I. | Upper 95% C.I. |
| Covid-19 Peritraumatic Distress Index (CPDI) |  | 58.99 | 0.000 | 29.14 | 88.83 |
| Normal range (4-27) | 412 (38.9%) |  |  |  |  |
| Mildly to moderately distressed (28-51) | 497 (47.0%) |  |  |  |  |
| Severely distressed (52-100) | 149 (14.1%) |  |  |  |  |
| Gender |  |  |  |  |  |
| Female | 569 (53.8%) | reference group |  |  |  |
| Male | 489 (46.2%) | -3.62 | 0.000 | -5.61 | -1.63 |
| Age |  |  |  |  |  |
| 18–25 years old | 137 (12.9%) | 0.81 | 0.170 | -0.35 | 1.98 |
| 26–35 years old | 364 (34.4%) |  |  |  |  |
| 36–45 years old | 358 (33.8%) |  |  |  |  |
| 46–55 years old | 161 (15.2%) |  |  |  |  |
| 56–65 years old | 33 (3.1%) |  |  |  |  |
| Over 65 years old | 5 (0.05%) |  |  |  |  |
| Education level |  |  |  |  |  |
| Junior high school or less | 24 (2.3%) | 0.37 | 0.286 | -0.31 | 1.05 |
| Senior high school | 30 (2.8%) |  |  |  |  |
| High school diploma | 122 (11.5%) |  |  |  |  |
| 2-year college degree | 97 (9.2%) |  |  |  |  |
| Bachelor degree | 411 (38.8%) |  |  |  |  |
| Master degree | 258 (24.4%) |  |  |  |  |
| PhD | 116 (11.0%) |  |  |  |  |
| Number of children |  |  |  |  |  |
| 0 | 422 (39.9%) | -1.07 | 0.048 | -2.14 | -0.01 |
| 1 | 250 (23.6%) |  |  |  |  |
| 2 | 305 (28.8%) |  |  |  |  |
| 3 | 61 (5.8%) |  |  |  |  |
| 4 or more | 20 (1.9%) |  |  |  |  |
| Infected by Covid-19 |  |  |  |  |  |
| Unsure | 182 (17.2%) | reference group |  |  |  |
| No | 869 (82.1%) | -8.91 | 0.000 | -11.26 | -6.57 |
| Yes | 7 (0.07%) | -9.43 | 0.090 | -20.34 | 1.48 |
| Exercise hours per day in the past week |  |  |  |  |  |
| 0 hours | 666 (62.9%) | -2.14 | 0.000 | -3.23 | -1.06 |
| 1 hour | 320 (30.2%) |  |  |  |  |
| 2 hours | 46 (4.3%) |  |  |  |  |
| 3 hours | 12 (1.1%) |  |  |  |  |
| 4 hours | 4 (0.04%) |  |  |  |  |
| 5 hours or more | 10 (0.09%) |  |  |  |  |
| Work situation |  |  |  |  |  |
| Stopped working due to Covid-19 | 281 (26.6%) | reference group |  |  |  |
| Worked at office | 179 (16.9%) | -2.63 | 0.059 | -5.36 | 0.10 |
| Worked from home | 408 (38.6%) | -2.87 | 0.012 | -5.11 | -0.63 |
| No work before and during outbreak | 190 (18.0%) | -3.44 | 0.023 | -6.42 | -0.47 |
| Employment status |  |  |  |  |  |
| Unemployed | 140 (13.2%) | reference group |  |  |  |
| Employed | 757 (71.5%) | -3.89 | 0.020 | -7.17 | -0.60 |
| Student | 111 (10.5%) | -5.29 | 0.020 | -9.74 | -0.85 |
| Retired | 50 (4.7%) | -5.19 | 0.071 | -10.83 | 0.45 |

The participants reported their distress level in the past week using CPDI, which had a Cronbach’s alpha of 0.91. The mean (SD) score of CPDI in the sample was 34.54 (14.92), higher than the CPDI of 23.65 (15.45) reported in China from January 31 to February 10, 2020^5^. The difference in the mean values between Iranian and Chinese samples is 10.9 (*t*=22.7; *p*<0.0001; 95% *CI*: 10.0 to 11.8). Based on the cut-off values of distress in CPDI, respectively 47.0% and 14.1% of the Iranian adults experienced mild to moderate and severe psychological distress, compared to 29.3% and 5.1% respectively in the Chinese sample^5^.

To analyze the identifiers (predictors) of CPDI, we used least square regression with the statistical significance of p□<□0.05 (Table 1 reports the regression coefficient, p-value, and the lower and upper 95% C.I.). Females experienced more distress than males (*β*=-3.62, *p*=0.000, 95% *CI* −5.61 to −1.63), similar to prior findings from China^5^. Participants’ age (*β*=0.81, *p*=0.170, 95% *CI* −0.35 to 1.98) and education level (*β*=0.37, *p*=0.286, 95% *CI* −0.31 to 1.05) did not predict distress as they did in the Chinese sample^3^. Adults with more children felt less distress (*β*=-1.07, *p*=0.048, 95% *CI* −2.14 to −0.01). Adults who were unsure whether they had Covid-19 reported higher distress than those who reported being free from the disease (*β*=-8.91, *p*=0.000, 95% *CI* −11.26 to −6.57). There is no significant difference between those who reported having the disease and the rest, possibly due to the very small number (7) of confirmed Covid-19 cases in our sample. Adults who exercised more reported lower distress (*β*=-2.14, *p*=0.000, 95% *CI* −3.23 to −1.06). Adults who were not working due to the Covid-19 pandemic reported higher distress than those who worked from home (*β*=-2.87, *p*=0.012, 95% *CI* - 5.11 to −0.63), at the office (*β*=-2.63, *p*=0.059, 95% *CI* −5.36 to 0.10), or who were not working even before Covid-19 (*β*=-3.44, *p*=0.023, 95% *CI* −6.42 to −0.47). The unemployed reported higher distress than the employed (*β*=-3.89, *p*=0.020, 95% *CI* −7.17 to −0.60) and students (*β*=-5.29, *p*=0.020, 95% *CI* −9.74 to −0.85). There is no significant difference in distress between the retired and the rest of the sample.

## Discussion

Although Covid-19 is expected to wreak havoc on the mental health of the public^1–3^, there is little evidence of it, especially outside of China. Our findings show that Iranian adults experienced significantly more distress (mean CPDI=34.5) than Chinese adults (mean CPDI=23.7). In late March 2020, a staggering 47.0% and 14.1% of Iranians in our sample reported mild to moderate and severe distress – also more severe than in China^5^.

Our findings suggest that the predictors of distress are likely to vary across countries. While gender and exercise hours predicted distress in both Iran and China^3,5^, age and education predicted distress in China but not in Iran^5^. It is worth noting another study also found age was not a significant predictor of distress among working adults in China during Covid-19^3^. Moreover, Iranian adults who worked from home, at the office, or had not worked during and before Covid-19 all reported lower distress that those who were not working due to the Covid-19 pandemic. In comparison, in China, only individuals who worked at an office reported significantly lower distress than those who were not working due to Covid-19^3^.

The differences in the predictors of distress during the Covid-19 pandemic across Iran and China are understandable, as different countries vary in their medical systems, the availability of personal protective equipment (PPE), cultures, labor and employment conditions, the policies of lockdown, the ease of working from home and maintaining a living in a pandemic, and the information in both mainstream and social media, to name just a few. The results therefore suggest we need to identify useful predictors of mental health in individual countries during the Covid-19 pandemic.

Lastly, individuals who were unsure whether they had Covid-19 reported higher distress than individuals who reported not having the disease, suggesting we need to pay attention to those individuals who might be anxious without knowing their specific Covid-19 status, as people can be asymptomatic or take time to develop symptoms. One solution is to test such individuals if conditions permit, as otherwise these individuals can experience high distress.

The study has certain limitations. First, we conducted this study using an observational cross-sectional survey, so our findings are predictive instead of causal. Second, our sample is not nationally representative, as our focus was to identify for policymakers and potential caregivers who in the population might need more help. Third, even though our sample contained adults who reported having Covid-19, the number of infected cases in our sample is too small, and moreover we suspect they would belong to the majority of Covid-19 infected cases that have mild symptoms, as people with more severe symptoms would not be able to answer our survey. Lastly, the scale has been translated from the Chinese version, which was newly developed in China during Covid-19, and further validation studies are needed on the scale.

In sum, this paper provides the first empirical evidence of the level of distress of Iranian adults during the Covid-19 pandemic. The results suggest adults in Iran are experiencing more distress than adults in China, with level of distress predicted by different factors, suggesting future research needs to examine mental health and the predictors in individual countries to effectively identify and screen those who are more susceptible to mental health issues during the Covid-19 pandemic.

## Data Availability

available upon request

## Acknowledgement

None.

## Contributors

**A.A.J**.: Investigation (data collection), Resources, Conceptualization, Writing – Review & Editing

**M.M.D:** Investigation (data collection)

**A.N.M**.: Investigation (data collection)

**J.L**.: Writing – Review & Editing; Funding acquisition

**S.X.Z**.: Conceptualization, Investigation, Methodology, Formal analysis, Visualization, Writing – Original, Writing – Review & Editing, Supervision

---

**Appendix A.** The Covid-19 Peritraumatic Distress Index (CPDI) in English. Please select the frequency of the below activities in the last week.

|  | Never | Occasionally | Sometimes | Often | Most of the time |
| --- | --- | --- | --- | --- | --- |
| 1. Compared to usual, I feel more nervous and anxious | 0 | 1 | 2 | 3 | 4 |
| 2. I feel insecure and bought a lot of goods, such as medications, sanitizer, gloves, masks, and/or other home supplies | 0 | 1 | 2 | 3 | 4 |
| 3. I can't stop myself from imagining myself or my family being infected and feel terrified and anxious about it | 0 | 1 | 2 | 3 | 4 |
| 4. I feel empty and helpless no matter what I do | 0 | 1 | 2 | 3 | 4 |
| 5. I feel sympathetic to the Covid-19 patients and their families. I feel sad about them. | 0 | 1 | 2 | 3 | 4 |
| 6. I feel helpless and angry about people around me, such as the governors and media | 0 | 1 | 2 | 3 | 4 |
| 7. I am losing faith in the people around me | 0 | 1 | 2 | 3 | 4 |
| 8. I collect information about Covid-19 all day. Even if it's not necessary, I can't stop myself | 0 | 1 | 2 | 3 | 4 |
| 9. I will believe the Covid-19 information from all sources without any evaluation | 0 | 1 | 2 | 3 | 4 |
| 10. I would rather believe in negative news about Covid-19 and be skeptical about the good news | 0 | 1 | 2 | 3 | 4 |
| 11. I am constantly sharing news about Covid-19 (mostly negative news) | 0 | 1 | 2 | 3 | 4 |
| 12. I avoid watching Covid-19 news, since I am too scared to do so | 0 | 1 | 2 | 3 | 4 |
| 13. I am more irritable and have frequent conflicts with my family | 0 | 1 | 2 | 3 | 4 |
| 14. I feel tired and sometimes even exhausted | 0 | 1 | 2 | 3 | 4 |
| 15. Due to feelings of anxiety, my reactions are becoming sluggish. | 0 | 1 | 2 | 3 | 4 |
| 16. I find it hard to concentrate | 0 | 1 | 2 | 3 | 4 |
| 17. I find it hard to make any decisions | 0 | 1 | 2 | 3 | 4 |
| 18. During this Covid-19 outbreak, I often feel dizzy, have back pain, or chest discomfort | 0 | 1 | 2 | 3 | 4 |
| 19. During this Covid-19 outbreak, I often feel stomach pain, bloating, or other stomach discomforts | 0 | 1 | 2 | 3 | 4 |
| 20. I feel uncomfortable when communicating with others | 0 | 1 | 2 | 3 | 4 |
| 21. Recently, I rarely talk to my family | 0 | 1 | 2 | 3 | 4 |
| 22. I cannot sleep well. I always dream about myself or my family being infected by coronavirus | 0 | 1 | 2 | 3 | 4 |
| 23. I lost my appetite | 0 | 1 | 2 | 3 | 4 |
| 24. I have constipation or frequent urination | 0 | 1 | 2 | 3 | 4 |

**Note:** CPDI = sum of the score of each question +4.
CPDI > 52: severe distress; CPDI within 28-51: mild/moderate distress; CPDI < 28: Normal;

**Appendix B.** The Covid-19 Peritraumatic Distress Index (CPDI) in Persian

پرسشنامه اثرات روحی روانی ویروس کرونا
| همیشه | غالب اوقات | بعضی اوقات | به ندرت | هرگز |  |
| --- | --- | --- | --- | --- | --- |
| 4 | 3 | 2 | 1 | 0 | 1- در مقایسه با روزهای معمولی گذشته ، این روزها احساس اضطراب و نگرانی بیشتری می‌کنم. |
| 4 | 3 | 2 | 1 | 0 | 2- احساس امنیت نمی‌کنم به همین دلیل کالاهای زیادی از قبیل داروها، ضدعفونی کننده، دستکش، ماسک و/ یا سایر لوازم ضروری را خریداری کرده ام. |
| 4 | 3 | 2 | 1 | 0 | 3- احساس می‌کنم خودم یا اعضای خانواده ام درگیر این بیماری خواهیم شد و نسبت به این موضوع وحشت زده و مضطرب هستم. |
| 4 | 3 | 2 | 1 | 0 | 4- بدون توجه به آنچه که در حال حاضر انجام می‌دهم بطور کلی احساس یاس و ناامیدی می‌کنم. |
| 4 | 3 | 2 | 1 | 0 | 5- من نسبت به مبتلایان به ویروس کرونا و خانواده های آنها احساس همدردی می‌کنم و بخاطر آنها ناراحت هستم. |
| 4 | 3 | 2 | 1 | 0 | 6- من نسبت به نحوه عملکرد در مورد ویروس کرونا، دولت و روزنامه ها احساس ناامیدی دارم. |
| 4 | 3 | 2 | 1 | 0 | 7- من امیدم را نسبت به سلامتی اطرافیانم در آینده نزدیک از دست می‌دهم. |
| 4 | 3 | 2 | 1 | 0 | 8- من تمام اطلاعات روز در مورد ویروس کرونا را جمع اوری می‌کنم. حتی اگر این اطلاعات لازم نباشند، نمی‌توانم جلوی خودم را بگیرم. |
| 4 | 3 | 2 | 1 | 0 | 9- من اطلاعات راجع به ویروس کرونا را از منابع مختلف و بدون ارزیابی، باور می‌کنم. |
| 4 | 3 | 2 | 1 | 0 | 10- اخبار منفی درباره ویروس کرونا را بیشتر باور می‌کنم تا خبرهای خوب و مثبت. |
| 4 | 3 | 2 | 1 | 0 | 11- من دائماً در مورد اخبار ویروس کرونا (عمدتاً اخبار منفی) با دوستان، آشنایان و همکاران صحبت می‌کنم. |
| 4 | 3 | 2 | 1 | 0 | 12- من از تماشای اخبار ویروس کرونا پرهیز می‌کنم ، زیرا من از این کار خیلی می‌ترسم. |
| 4 | 3 | 2 | 1 | 0 | 13- این روزها من خیلی حساس و تحریک پذیر شده ام و جر و بحث های مکرری با خانواده دارم. |
| 4 | 3 | 2 | 1 | 0 | 14- احساس خستگی و ناامیدی زیادی می‌کنم . |
| 4 | 3 | 2 | 1 | 0 | 15- به دلیل احساس اضطراب زیاد، واکنش های من به محیط اطراف کند شده است. |
| 4 | 3 | 2 | 1 | 0 | 16- تمرکز کردن برایم خیلی دشوار شده است. |
| 4 | 3 | 2 | 1 | 0 | 17- تصمیم گیری برایم خیلی سخت شده است. |
| 4 | 3 | 2 | 1 | 0 | 18- طی دوره وجود ویروس کرونا در کشور، من اغلب احساس سرگیجه ، کمردرد یا ناراحتی در قفسه سینه می‌کنم. |
| 4 | 3 | 2 | 1 | 0 | 19- طی دوره وجود ویروس کرونا در کشور، من اغلب احساس درد معده ، نفخ یا سایر ناراحتی های معده می‌کنم. |
| 4 | 3 | 2 | 1 | 0 | 20- این روزها هنگام برقراری ارتباط با دیگران احساس ناراحتی می‌کنم. |
| 4 | 3 | 2 | 1 | 0 | 21- این روزها، به ندرت با خانواده ام صحبت می‌کنم کم حرف شده ام. |
| 4 | 3 | 2 | 1 | 0 | 22- این روزها نمی‌توانم خوب بخوابم. همیشه خواب می‌بینم که خودم یا خانواده ام به ویروس کرونا آلوده شده ایم. |
| 4 | 3 | 2 | 1 | 0 | 23- اشتهايم را از دست داده ام. |
| 4 | 3 | 2 | 1 | 0 | 24- این روزها دچار یبوست یا تکرر ادرار شده ام. |

## References

1 Xiang Y-T, Yang Y, Li W, et al. Timely mental health care for the 2019 novel coronavirus outbreak is urgently needed. The Lancet Psychiatry 2020; 7: 228–9.

2 Bao Y, Sun Y, Meng S, Shi J, Lu L. 2019-nCoV epidemic: address mental health care to empower society. Lancet 2020; 395: e37–8.

3 Zhang SX, Wang Y, Rauch A, Wei F. Unprecedented disruptions of lives and work: Health, distress and life satisfaction of working adults in China one month into the COVID-19 outbreak. Psychiatry Res 2020; published online March 30. DOI:10.1101/2020.03.13.20034496.

4 Gao J, Zheng P, Jia Y, et al. Mental Health Problems and Social Media Exposure During COVID-19 Outbreak. SSRN Electron J 2020; published online Feb 17. DOI:10.2139/ssrn.3541120.

5 Qiu J, Shen B, Zhao M, Wang Z, Xie B, Yifeng Xu. A nationwide survey of psychological distress among Chinese people in the COVID-19 epidemic: implications and policy recommendations. Gen Psychiatry 2020; 33: 1–6.

6 Zandifar A, Badrfam R. Iranian mental health during the COVID-19 epidemic. Asian J Psychiatr 2020; 51: 101990.

7 Takian A, Raoofi A, Kazempour-Ardebili S. COVID-19 battle during the toughest sanctions against Iran. Lancet 2020; 2019: 30668.

